# Precision recruitment for high-risk participants in a COVID-19 research study

**DOI:** 10.1101/2022.03.03.22271504

**Authors:** Aziz M. Mezlini, Eamon Caddigan, Allison Shapiro, Ernesto Ramirez, Helena M. Kondow-McConaghy, Justin Yang, Kerry DeMarco, Pejman Naraghi-Arani, Luca Foschini

## Abstract

Studies for developing diagnostics and treatments for infectious diseases usually require observing the onset of infection during the study period. However, when the infection base rate incidence is low, the cohort size required to measure an effect becomes large, and recruitment becomes costly and prolonged. We describe an approach for reducing recruiting time and resources in a COVID-19 study by targeting recruitment to high-risk individuals. Our approach is based on direct and longitudinal connection with research participants and computes individual risk scores from individually permissioned data about socioeconomic and behavioural data, in combination with predicted local prevalence data. When we used these scores to recruit a balanced cohort of participants for a COVID-19 detection study, we obtained a 4–7-fold greater COVID-19 infection incidence compared with similar real-world study cohorts.

---

The costs of recruiting large numbers of participants for clinical studies can be high. In addition, the power of clinical trials can depend on the number of ‘rare events’ observed (such as COVID-19 infections), which often takes long periods to accrue. Given these factors, efforts have been attempted to reduce costs, increase the power of analyses, and shorten study periods by targeting participants at higher risk (those who were more exposed to infection) during the recruitment process^1^.

We present an enrichment approach based on direct connection with members of the Evidation health and research platform^2^. This application (Evidation Health, Inc., San Mateo, CA) is a reward platform that encourages users to develop healthy habits—such as walking, meditating, and logging meals—and provides incentives for them to participate in research by completing surveys and sharing their data from commercial-grade wearable sensors^3,4^. For example, the application has been used since 2017 for voluntary monitoring of annual waves of influenza cases^5^. We therefore had access to a large pool of potential study participants we could easily survey.

We leveraged our longitudinal prerecruitment relationship with members of the Evidation platform and applied machine learning (ML) modelling of COVID-19 risk to recruit a high-risk study population. The ML model quantified risk using participants’ locations and responses to questions about their occupations and behaviour, given that these features are likely to affect exposure level to the virus. The initial training phase for the model was 49 days, during which we followed more than 100,000 members to characterize those who contracted COVID-19.

We measured enrichment in terms of the incidence of COVID-19 infection obtained using this enhanced selection process compared with the incidence of COVID-19 infections in the control groups of three COVID-19 vaccine trials^6-8^ and in another cohort generated via the Evidation platform but not using a precision recruitment approach^9^. Because each study had different enrolment dates and demographics, we normalized the incidence in each study by matching the dates and demographics of the comparator study to the US incidence.

Specifically, for this study, we launched an initial Risk of Occupational Exposure to COVID-19 deep-labelling survey on June 15, 2020. This survey, which collected demographic, socio-economic, and behavioural data on potential high-risk populations, had received 128,629 responses at the time of this analysis. Short follow-up surveys were sent to respondents to the Risk of Occupational Exposure survey who had indicated they had not been diagnosed with, nor experienced symptoms related to, COVID-19 as of August 3, 2020. The follow-up survey was used to determine if any individuals had received a positive diagnosis (and date of diagnosis) in the interim period (approximately 2 months) since completing the initial survey. Of the 94,7000 who were sent the follow-up survey, 66,040 responded, and 514 (0.8%) indicated they had received a COVID-19 diagnosis in the interim.

We then created an ML model using labelling responses from the initial survey, which performed better than chance at identifying who would receive a subsequent diagnosis. This model incorporated predictions of COVID-19 local prevalence (using generalized additive models [GAMs]) as a variable, along with socioeconomic and behavioural data from the initial survey. The model was trained only on participants who responded to the second survey, with the outcome variable being whether they had contracted COVID-19 during the 49-day follow-up period. The model was trained using random forests. See the Supplemental Material for a detailed description of the modelling process.

The trained model was then used to calculate a risk score for each person who had responded to the initial survey. Persons with the highest risk scores were primarily targeted for recruitment and were selected to generate a dataset with balanced demographics (such as age, sex, and ethnicity).

To compare our findings with enrolment in the other studies, we first calculated the incidence rate as the number of confirmed COVID-19 cases per 1000 person-years of follow-up for our cohort and for each comparison cohort. Only the person-days at risk of contracting COVID-19 were considered, and all days occurring after vaccination or contracting COVID-19 were excluded. With this method, breakthrough infections did not end up being counted in our tally, therefore providing a conservative estimate.

To calculate the US-matched incidence for our cohort, we used individual-level data from the Centers for Disease Control and Prevention (CDC) describing all confirmed COVID-19 cases in the U.S. (Of note, these data likely underestimate the true number of cases.) We aggregated counts by date and by demographic group (sex by age group).

For each comparator study, we calculated the US incidence of COVID-19 during the study period for each demographic group, using 2019 US Census data for the size of each demographic group in the U.S.^10^ To calculate the final US-matched incidence, we measured the proportion of each demographic group in the comparator study, and then took the weighted average of the US incidences across demographic groups (weighted by the proportions of these groups in the comparator study). By dividing each study incidence by the US-matched incidence, we ensured that our findings were not biased by differences in the study period or demographics. The 95% confidence intervals for the incidence in each study and the ratios were calculated using the exact method (Poisson distribution).

From the candidates with the highest risk scores, we recruited a demographically balanced dataset of 840 participants and followed them from November 5, 2020 to April 15th, 2021. The total follow-up time to reported infection or vaccination was 141.2 person-years, and 104 participants (12.3%) developed confirmed COVID-19 infection.

Comparing our model with recruitment in other studies, we observed 4- to 7-fold greater detection of COVID-19 cases after accounting for differences in study periods and the numbers of COVID-19 cases in the U.S. at those times (Figure 1). See Supplemental Table 1 for the data listings for each study included in the analysis.

**Figure 1.**
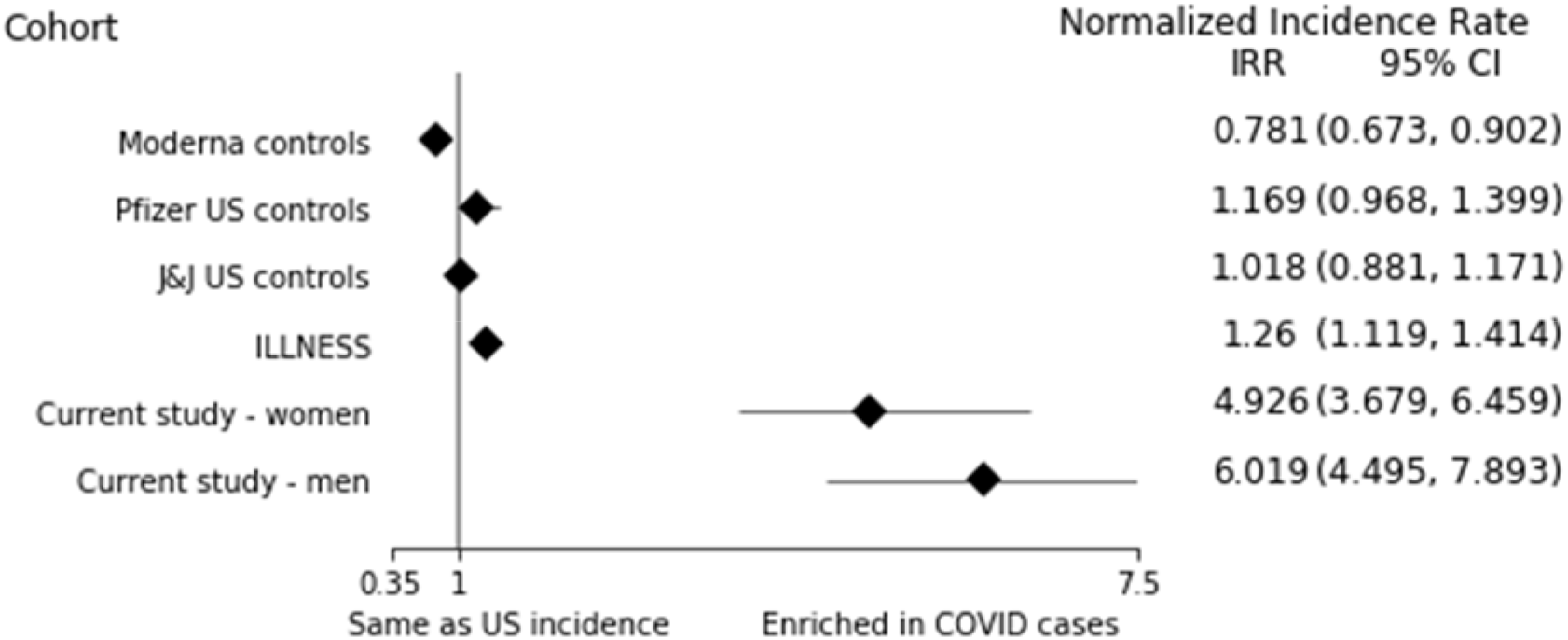
Covid-19 incidence rate in each cohort normalized by the US incidence rate matched for time, age, and sex.

Figure 2 shows the variables most important to the prediction of COVID-19 infection. The top features related to the number of potentially risky contacts (household size and residential situation), location (living in a city with numerous COVID-19 cases at the time of recruitment) and working in a risky occupation (healthcare workers). See the Supplement for more details about these features. In contrast, the features that were least important were related to non-healthcare work settings (hospitality, public transit, agriculture, self-employed, etc.)

**Figure 2.**
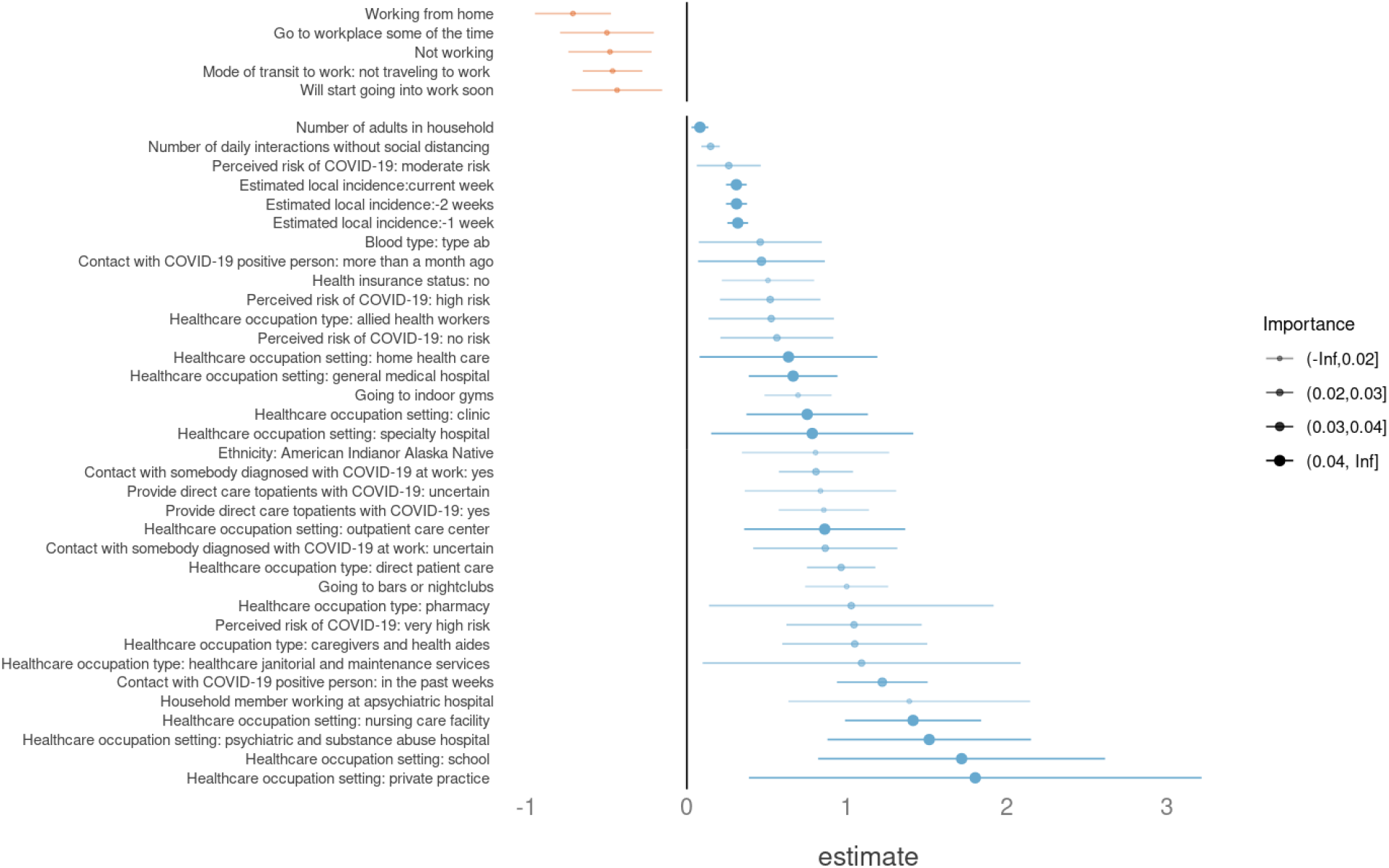
The most important 20 groups of predictors of risk in our model ranked by random forest feature importance. Sub-predictors from the same group (e.g Healthcare occupation) have been separated and rearranged for visual clarity. For each variable, we show the direction of effect based on the coefficient of a logistic regression and its confidence interval. Positive coefficients that are statistically significant (risk increasing) are in blue, and negative ones are in orange. For categorical variables, we show the coefficient corresponding to each level vs. the reference level (none/no answer). More detailed descriptions of each factor and variable are available in the Supplement.

Although our risk model showed up to a 7-fold increase in recruitment possibility, we can improve it further through including additional variables that might be relevant, such as information about contacts with other people, especially with children of school age. We also could improve the training process and GAM models.

The current analysis has several limitations of note. For some of the datasets (Moderna, J&J, Pfizer), we had access only to the limited publicly available data. For example, we had no access to individual data about each participant’s start and end date in the trial. Therefore, when computing the matched US incidence for each of these trials (matched in time, age, and sex), we used the overall study start and end date of each trial. This does not consider variability in when each participant started and ended the trial. If a study had very gradual or slow enrolment, its incidence might have differed substantially from the US-matched incidence over the duration of the study.

Additionally, some studies tallied only events happening 14 days after the second vaccination dose/placebo. Given that we did not have individual-level data for when each participant received their second dose, we could not account for that variability. Instead, as shown in Supplementary Table 1, we recalculated the US-matched incidences after removing the initial days from each study corresponding to the length of its vaccination protocol plus 14 days (instead of using the full length of the study). This attempt to count from a later start date resulted in a higher US-matched incidence and therefore even lower comparative incidence ratios for the other vaccine trials. Thus, our current estimate of enrichment compared with other studies is a lower bound on the real enrichment. Overall, our comparisons to US-matched incidences are imperfect (because of lack of individual-level data) but consistent across studies.

In this paper, we compared our precision recruitment approach to other internal (Illness) and external (Moderna, J&J, Pfizer) trials while taking into account that these trials happened at different times and with different sample demographics. We used calculations of matched US incidences (in time, age, and sex) for each trial to be able to properly make those comparisons. The comparisons showed a substantial enrichment factor of 4–7 times the incidence of COVID-19 infections. This means that our precision recruitment approach could be applied to reduce the needed sample size of future trials by this factor, or shorten the trials’ duration by the same factor. Beyond the benefit of reducing the cost of clinical trials, this precision recruitment approach could be of great utility in situations of emergency such as during a pandemic.

Whether the findings obtained on the recruited population would be generalizable to the full population is not guaranteed in general when using a technique for precision recruitment as described. For a vaccine trial this should be true, given that our population was not biologically different in demographics or comorbidities. The difference with the full population relates only to greater physical exposure to COVID-19 (through location, occupation, and behaviour) rather than difference in biological susceptibility to COVID-19. Therefore, we do not expect any conclusion made regarding vaccine efficacy to be affected by our precision recruitment procedure, and results should be generalisable to other populations.

## Data Availability

The datasets analysed in this study are not publicly available but can be shared for scientific collaboration by contacting the corresponding author.

## Acknowledgements

This manuscript is not an endorsement of any technology or platform. This project was funded in whole or in part with federal funds from the Department of Health and Human Services, Office of the Assistant Secretary for Preparedness and Response, Biomedical Advanced Research and Development Authority, under contract number 75A50120C00091. Additional support was provided by an appointment to the Biomedical Advanced Research and Development Authority (BARDA) Research Participation Program administered by the Oak Ridge Institute for Science and Education (ORISE) through an interagency agreement between the U.S. Department of Energy (DOE) and the U.S. Department of Health and Human Services (HHS). Opinions expressed in this paper are the authors’ and do not necessarily reflect the views of HHS, BARDA, DOE, or ORAU/ORISE.

## Author Contributions

A.M.: data analysis, comparison with other studies, writing.

E.C.: risk model, features importance.

P.N.A.: selection criteria for recruitment of study participants.

