## Supplemental Material for "Precision recruitment for high-risk participants in a COVID-19 research study"

**Covid Hotspot Model**

We built a simple Covid Hotspot prediction model using county-level incidence rates reported by the *New York Times*^1^. For starting Week 4 through 29, a three-dimensional general additive model (GAM) was fitted to 10 weeks of incidence rate data, with the latitude and longitude of the county centroid and week number as predictors. The mean squared error (MSE) of this model’s predictions for the 2 weeks after the training period (i.e., Weeks 14 and 15 through 31 and 32) was calculated across the following hyperparameters: smoother for the longitude and latitude (tensor product or cubic regression with shrinkage) and the rank of the smooths (4, 6, or 8). Supplemental Figure 1 shows the performance of the GAM.

**Supplemental Figure 1. GAM performance. Concordance with real COVID-19 hotspots at the time of recruitment.**

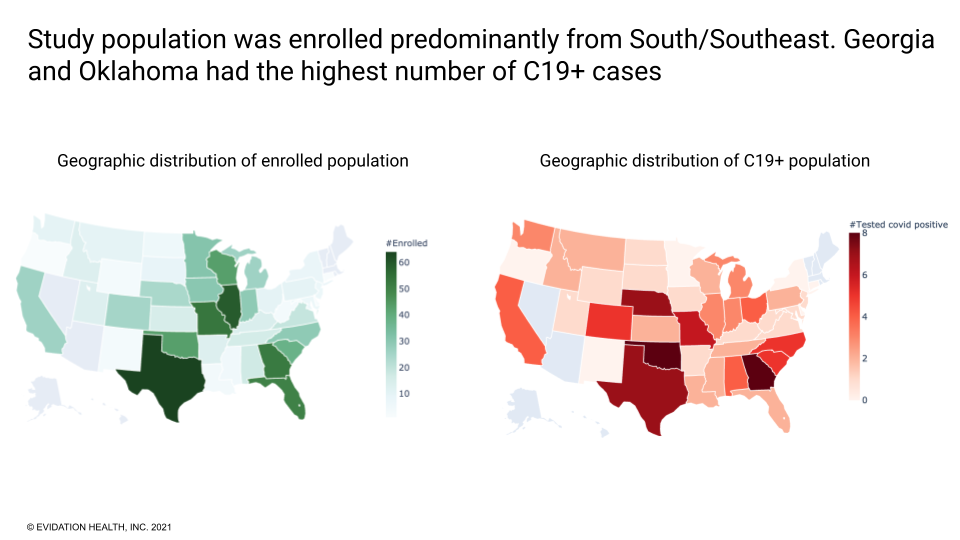

The best-performing model used a rank-6 temporal product smoother for the spatial components, and a rank-4 cubic regression spline with shrinkage for week number. These hyperparameters were then used to build a series of models that predicted the incidence rate for each ZIP code tabulation area (ZCTA) and week number of the response period using the county-level incidence rate for the preceding 10 weeks.

For each observation in our COVID-19 labeling dataset, three predictors were added using these models: the *predicted* COVID-19 incidence rate for the respondents’ home ZCTA at the week of diagnosis, and the predictions for the preceding 2 weeks. Respondents who did not report a COVID-19 diagnosis were assigned a random “week of diagnosis” for this purpose.

**COVID-19 Risk Model**

We used the data collected about COVID-19 cases in the follow-up period between the initial survey and the second survey. The labels used for training the risk model is whether the considered participant had COVID-19 in that follow-up period (June 15, 2020 to August 3, 2020).

The features used consisted of all the information obtained in survey one (socioeconomic, behavioral, occupation) plus the predictions from the Hotspot prediction model for the participant location.

We used Random Forests ([Spark’s random forest classifier](http://spark.apache.org/docs/latest/ml-classification-regression.html#random-forest-classifier), via tidymodels/parsnip). After hyperparameter tuning with 5 inner folds, we used the number of trees equal to 1024 to train the model. The model was evaluated by choosing a test week and training a classifier on the preceding weeks across participants (this way, nothing was “contaminated” by information from the future).

After training, we applied the model on all participants who responded to the initial survey, using the GAM predictions for the expected recruitment date, in order to calculate risk scores at the future time of recruitment.

During recruitment, we only selected participants with no previous COVID-19 infection reported. We used the risk scores to separately enroll the highest risk men and women. Some manual tweaking was involved to ensure a balanced representation across ages and ethnicities among the high-risk candidates selected.

**Important variables definitions**

Supplemental Figure 2 shows the most important features reflecting individuals COVID-19 risk calculation. Here we describe the details of our features:

- **Adult Household Size:** Answer to the following question, “How many adults (age 18+) currently live in your household (including yourself)? Include roommates, or if you are living in a group home, the total number of adults you share common living spaces with.”
- **Estimated local incidence:** We built a series of hotspot prediction models using the county-level prevalence data provided by the New York Times. We fit a three-dimensional GAM to prevalence values with latitude, longitude, and (week number) as predictors, for weeks 1...*N*, and then used this model to predict prevalence for week *N*+1. For each observation used to train the model, this model predicted prevalence rate for the associated ZIP code for the week number of the response and the preceding two weeks.
- **Health occupation setting:** Response to, “In which healthcare setting do you primarily work in?” This was a categorical feature that had the levels ranked for the highest coefficient to the lowest in Figure 2. All levels were compared to “none” (participants not working in a health occupation setting). The values that were significantly associated with a higher risk were: “nursing care facility”, “general medical hospital”, “psychiatric and substance abuse hospital”, “clinic”, “school”, “outpatient care center”, “private practice”, “specialty hospital”, and “home health care”. The values that showed no significant difference from zero are: “pharmacy”, “residential care except skilled”, “home dental office”, “other”, “veterinary”, and “work from home”.
- **Contact with somebody diagnosed with COVID-19:** Response to, “Have you been in prolonged close contact (i.e., within 6 feet for at least 10 minutes) with someone who was diagnosed with COVID-19?” This was a categorical feature with the following levels “no”, “yes in the past weeks”, “yes in the past month”, and “yes but more than a month ago”. We used the “no” answer as the reference and saw that the level “yes in the past weeks” was highly associated with risk (*P* <0.001). The other two answers had a lower coefficient with “yes but more than a month ago”, reaching statistical significance (*P* = 0.02).
- **Healthcare occupation type:** Response to, “Do you work in one of the following healthcare-related occupations?” This was a categorical variable with 17 possible levels. The reference chosen was “none”. Most levels had a positive coefficient, but because of the lower sample size for each level, only the following levels reached statistical significance: “healthcare workers providing direct patient care”, “caregivers and health aides”, “allied health workers”, “pharmacy”, and “healthcare janitorial and maintenance services”. “Morgue workers” was the level with the highest coefficient.
- **Perceived risk of COVID-19:** Response to, “What do you believe is your personal risk of getting COVID-19? Please think about your job, your usual activities, your close contacts etc.” This was a categorical variable with the following levels: “very high risk”, “high risk”, “no risk”, “moderate risk”, “low risk”, and “very low risk”. We used the “low risk” level as the reference. Compared with that group, the highest risk level was in participants who answered “very high risk” and “high risk”, followed by participants who answered “no risk”, then “moderate risk”. All these levels were associated with increased risk and were statistically significant. This might indicate that participants who perceived they had no risk of contracting COVID-19 did not take sufficient protective measures. The level “very low risk” was the only level associated with a lower risk than the reference (not statistically significant).
- **Current residential situation:** Response to, “What is your current residential situation?” This was a categorical variable with the following levels ranked from the highest coefficient to the lowest: “assisted living facility”, “dormitory”, “other”, “with family”, “mobile home or RV”, “shared home”, “duplex”, “townhome”, “apartment or condo”, and “single family home”. We used the “single family home” level as the reference. Only the answer “dormitory” was close to statistical significance (*P =* 0.05)
- **Contact with somebody diagnosed with COVID-19 at work:** Response to, “On an average work day, how many individuals do you closely interact with that do not follow social distancing recommendations (i.e., have a face-to-face conversation, help with a task and do not stay at least 6 feet apart)?” This had three possible answers: “yes”, “uncertain”, and “no”. We used the answer “no” as the reference. Both of the other answers were associated with a higher risk.
- **Blood type:** had the following levels ranked from highest to lowest risk: “AB”, “B”, “A”, “O”, and “uncertain”. We used the level “uncertain” as a reference. The level “AB” was associated with a *P* value of 0.02.
- **Physically going to work:** had the following levels ranked from the highest to the lowest risk: “often”, “no, but I will start physically going soon”, and “no, not working”, “some of the time”, “no”. We used the level “often” as the reference.
- **Mode of transit:** had the following levels ranked from the highest to the lowest risk: “carpool with others who are not members of my household”, “take public transportation”, “walk and or use personal transportation”, “drive alone or with members of my household”, “not traveling to work”, “other”, and “Uber”. We used the level “drive alone or with members of my household” as the reference. Compared with the reference, the level “not traveling to work” reached statistical significance with a *P* value <0.001.
- **Provide direct care to patients with COVID-19:** had the following levels ranked from the highest to the lowest risk: “yes”, “uncertain”, “no”. We used the “no” answer as the reference.
- **Insurance status:** had the following levels ranked from the highest to the lowest risk: “no”, “uncertain”, “yes”. We used the “yes” answer as the reference.

**Supplemental Figure 2. Features most important to individual COVID-19 risk calculations**

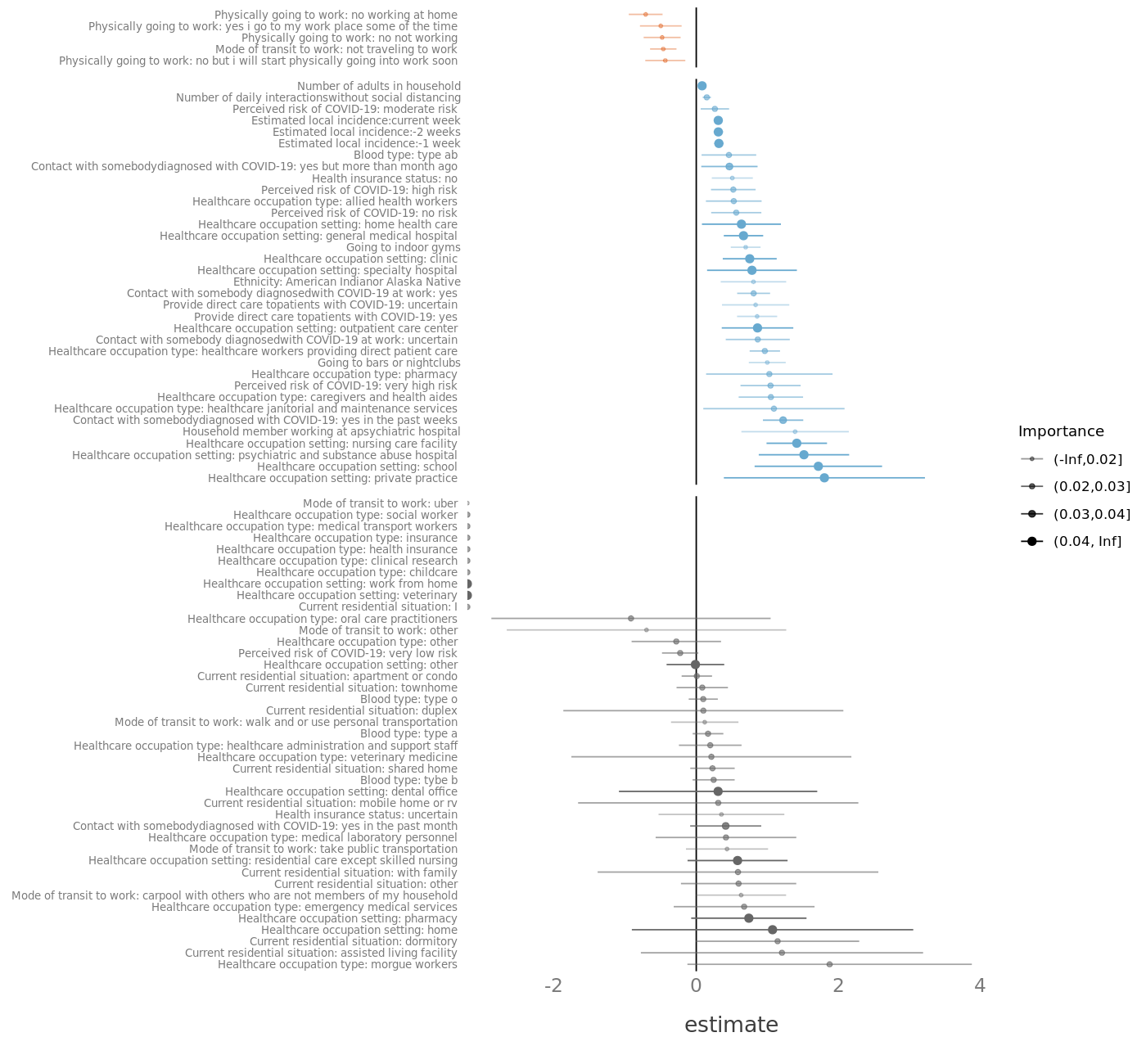

**Supplemental Table 1. Comparison of COVID incidence across trials**

|  | *N* | Study period | n/ person-year | Trial incidence | US-matched incidence | Incidence ratio |
| --- | --- | --- | --- | --- | --- | --- |
| Moderna controls^2^ | 14,073 | 2020/07/27 - 2020/11/25 | 185/3274 | 56.5 | 72.3 (88.6) | 0.78 (0.63) |
| Pfizer US controls^3^ | 13,506 | 2020/07/27 - 2020/11/14 | 119/1747 | 68.1 | 58.2 (64.1) | 1.16 (1.06) |
| J&J US controls^4^ | 9,086 | 2020/09/21 - 2021/01/22 | 196/1391 | 140.9 | 138.4 (150.1) | 1.01 (0.93) |
| ILLNESS study^5^ | 23,428 | 2021/02/09 - 2021/06/11 | 290/4131 | 70.2 | 55.7 | 1.26 |
| Current study - women | 452 | 2020/11/05 - 2021/04/15 | 52/76.5 | 679.5 | 138.0 | 4.92 |
| Current study - men | 388 | 2020/11/23 - 2021/04/15 | 52/64.7 | 804.0 | 133.5 | 6.02 |

We matched the incidence in each trial with the CDC-reported US incidence over the same duration of the trial, matched by age and sex. Incidence corresponds to the number of cases per 1000 person-years. The reported numbers correspond to COVID-19 onset 14 days after placebo vaccination #2 for J&J and Moderna, and 7 days after the second dose for Pfizer. For time-matched US incidence, the numbers in parentheses represent the US matched incidence calculated if we had removed the first 14 days of the J&J trial, the first 28+14 days for Moderna, and the first 21+7 days from Pfizer (the time between the two doses was 28 days for Moderna and 21 days for Pfizer).
